# Effectiveness of lower limb rehabilitation protocol using mobile health on quality of life, functional strength and functional capacity among knee osteoarthritis patients who were overweight and obese: A randomized controlled trial

**DOI:** 10.1101/2022.01.05.21268595

**Authors:** Muhammad Tariq Rafiq, Mohamad Shariff Abdul Hamid, Eliza Hafiz

**Author notes:** Emails: Muhammad Tariq Rafiq; Mohamad Shariff Abdul Hamid; Eliza Hafiz.

## Abstract

**Objective:** This study aimed to investigate the effectiveness of the lower limb rehabilitation protocol (LLRP) using mobile health (mHealth) on quality of life (QoL), functional strength, and functional capacity among knee OA patients who were overweight and obese.

**Materials and Methods:** In the current trial, 114 patients were recruited and randomized into either the rehabilitation group with mobile health (RGw-mHealth) receiving reminders by using mHealth to carry on the strengthening exercises of LLRP and instructions of daily care (IDC), the rehabilitation group without mobile health (RGwo-mHealth) following the strengthening exercises of LLRP and instructions of daily care (IDC) and control group (CG) only following the IDC for duration of 12-weeks. The reminders for using mHealth were provided two times a day for three days a week. Primary outcome measures were QoL assessed by the Western Ontario and McMaster Universities Osteoarthritis Index summary score, and functional strength by Five-Repetition Sit-To-Stand Test. Secondary outcome measure was functional capacity assessed by the Gait Speed Test. The assessments of QoL, functional strength, and functional capacity were taken at baseline and posttest after 12-weeks of intervention.

**Results:** After 12 weeks of intervention, patients in all three groups had statistically significant improvement in QoL within groups (p < 0.05). Furthermore, patients in the RGw-mHealth and RGwo-mHealth had statistically significant improvement in functional strength and walking gait speed within groups (p < 0.05). The pairwise between-group comparisons (Bonferroni post hoc test) of the mean changes in QoL, functional strength, and functional capacity at posttest assessments revealed that patients in the RGw-mHealth had statistically significant greater mean change in QoL, functional strength and functional capacity relative to both the RGwo-mHealth and CG (p < 0.001).

**Conclusion:** Improvement in QoL, functional strength, and functional capacity was larger among patients in the RGw-mHealth compared with the RGwo-mHealth or CG.

**TRIAL REGISTRATION:** **Chinese Clinical Trial Registry:** ChiCTR1900028600

**Date of registration:** 28-12-2019

**Registration Status:** Prospective

**URL:** http://www.chictr.org.cn

---

In the United States, doctor-diagnosed arthritis is a common chronic condition (1). Osteoarthritis (OA) is a degenerative disease in character and can lead to functional limitations and muscle weakness (2). A published article pointed out that OA is linked to joint wear and tear as well as inflammation of the synovial membrane (3). OA is a chronic disease that occurs in the knees, hips, hands, and spinal joints and causes pain, stiffness, and decreased range of motion (ROM) (4). Generally, populations do not prefer to participate in physical activity. Lack of time and different types of exercise were cited as major barriers (5, 6). OA causes a considerable burden on the quality of life (QoL) and medical treatment of patients (7). In 2015, Knee OA was the highest contributor among OA of the thirteenth leading cause of global disability (8) and it diminished QoL (9). The five-repetition sit-to-stand test (FRSST) test was used for the measurement of functional strength of the lower body. The FRSST measures the time taken to complete the repeated action of a stand five times from a sitting position as rapidly as possible (10). A person’s ability to cope with daily life activities is known as functional capacity (11).

Smart phone’s mobile health applications (mHealth apps) have the potential to play an important role in supporting personal health management (12). A current systematic review found that mHealth app users were more satisfied to manage their health than those of conventional care. The mHealth app users have resulted in a positive impact on health outcomes and health-related behaviors (13).

Exercise therapy is a safe and low-cost method for treating Knee OA that has been shown to delay disease progression and improve knee function (14). The Ottawa Panel found evidence to support the use of therapeutic exercises, especially strengthening exercise and general physical activity, for the improvement of functional characteristics in OA patients (15). A current systematic review on non-pharmacological interventions for treating symptoms of knee OA in overweight or obese patients resulted that the most effective intervention that showed improvement of knee function was strengthening exercise (16). Non-pharmacological interventions, primarily strengthening exercise and more recently strengthening exercises of the lower limbs in non-weight-bearing positions, are recommended as the first line of treatment among overweight or obese knee OA patients (17). The novelty of the current study could have been mediated by two factors, firstly it was provided as text messages and secondly the researchers designed a new lower limb rehabilitation protocol (LLRP) to treat overweight and obese knee OA patients. The training sessions of the LLRP are the strengthening exercises of the major muscle groups of the lower limbs in non-weight-bearing sitting and lying positions to reduce the mechanical load on the knee. However, there is insufficient evidence on the effectiveness of the LLRP by using mHealth on QoL, functional strength, and functional capacity among overweight or obese knee OA patients. Therefore, the objective of the current study was to investigate the effectiveness of the LLRP using mHealth on QoL, functional strength, and functional capacity among knee OA patients who were overweight and obese.

## MATERIALS AND METHODS

### Design and setting

The assessments of outcome measures of the current randomized controlled trial (RCT) were conducted in the Teaching Bay. The study was approved by the regional Ethical Committee with approval No: RAIC PESSI /Estt/2019/487 on date 28-08-2019 and the trial was registered in the Chinese Clinical Trial Registry with registration number:ChiCTR1900028600 on date 28-12-2019. The first patient in the trial was enrolled on the date 02-01-2020. Pre-defined questionnaire of inclusion and exclusion criteria was used for screening of the patients.

### Study patients

Patients diagnosed with knee OA who were overweight or obese from the urban community were screened. According to World Health Organization, the individuals who have BMI ≥ 25kg/m^2^ are known as overweight and individuals who have BMI ≥ 30kg/m^2^ are known as obese (18). The sample included males and females diagnosed with 2-mild or 3-moderate OA according to Kellgren and Lawrence radiographic scale (19). The anteroposterior and lateral view of plain radiography of the affected knee/s were performed in the standing position.

The inclusion criteria were as follows: both males and females, overweight and obese knee OA patients; age 45-60 years; residing in an urban community. The exclusion criteria were as follows: system lupus erythematosus, rheumatoid arthritis, spondyloarthropathies, sjogren’s syndrome, gout, scleroderma, infectious arthritis; history of metabolic, hormonal, orthopaedic, cardiovascular disease; spinal deformities; flat feet; previous surgery of knee/s of any cause or injection of knee/s for the last 6-months. The experimental procedures, risks, and benefits associated with the study were explained to all patients prior to providing written informed consent.

### Sample size

Sample size estimation was performed using the G^*^ Power 3.1.3 software. By assuming the medium effect size f = 0.70, setting α = 0.05, power (1-B) = 0.80, the total sample size estimated was 30 patients for each group. After considering 20% drop-out rates, the sample size of 114 patients for the three groups was decided.

### Patient’s recruitment and selection

Patients were recruited from the urban area using convenience sampling by active recruitment strategies through the political and welfare organizations. A study coordinator explained the criteria of knee OA patients as well as benefits of study participation to the political and welfare organizations. These organizations explained the benefits to the expected knee OA patients in the recruitment area via word of mouth and presented a list of expected knee OA patients to a study coordinator. The study coordinator prepared a list of potential patients based on the inclusion and exclusion criteria of the study and approved a final list of patients to contact. Then, the study coordinator called patients to attend a meeting in the Teaching Bay to further assess eligibility. Patients who were eligible and interested in participating completed a written informed consent for their participation in the study. Prior to the intervention, patients’ baseline measurements were completed

### Blinding and Allocation

The coordinators collecting data were independent individuals from the trials and were unaware of the group allocation. There were different coordinators at the baseline and post-test evaluation. Individuals performing the statistical analysis were kept blinded by labelling the groups with non-identifying terms (such as X and Y).

### Study Randomization

After completing the screening for eligibility, the selected patients were randomized into one of three groups: rehabilitation group with mobile health (RGw-mHealth), rehabilitation group without mobile health (RGwo-mHealth) or control group (CG) by a computer-generated number. Each group consisted of thirty-eight patients. All patients were also given a diary and asked to record the attendance of completion their interventions based on leaflets.

### Research procedures

#### Rehabilitation group without mobile health (RGwo-mHealth)

Patients in the RGwo-mHealth were taught on how to perform the strengthening exercises of LLRP by following the IDC. The contents of IDC were explained elsewhere (17). Patients were advised to continue performing the training sessions of LLRP 3-times a week for 12-weeks at home. These trainings included strengthening exercises for the lower limbs in non-weight-bearing sitting, or lying positions **(Table 1)** without putting a mechanical load on the knee.

**Table 1.**
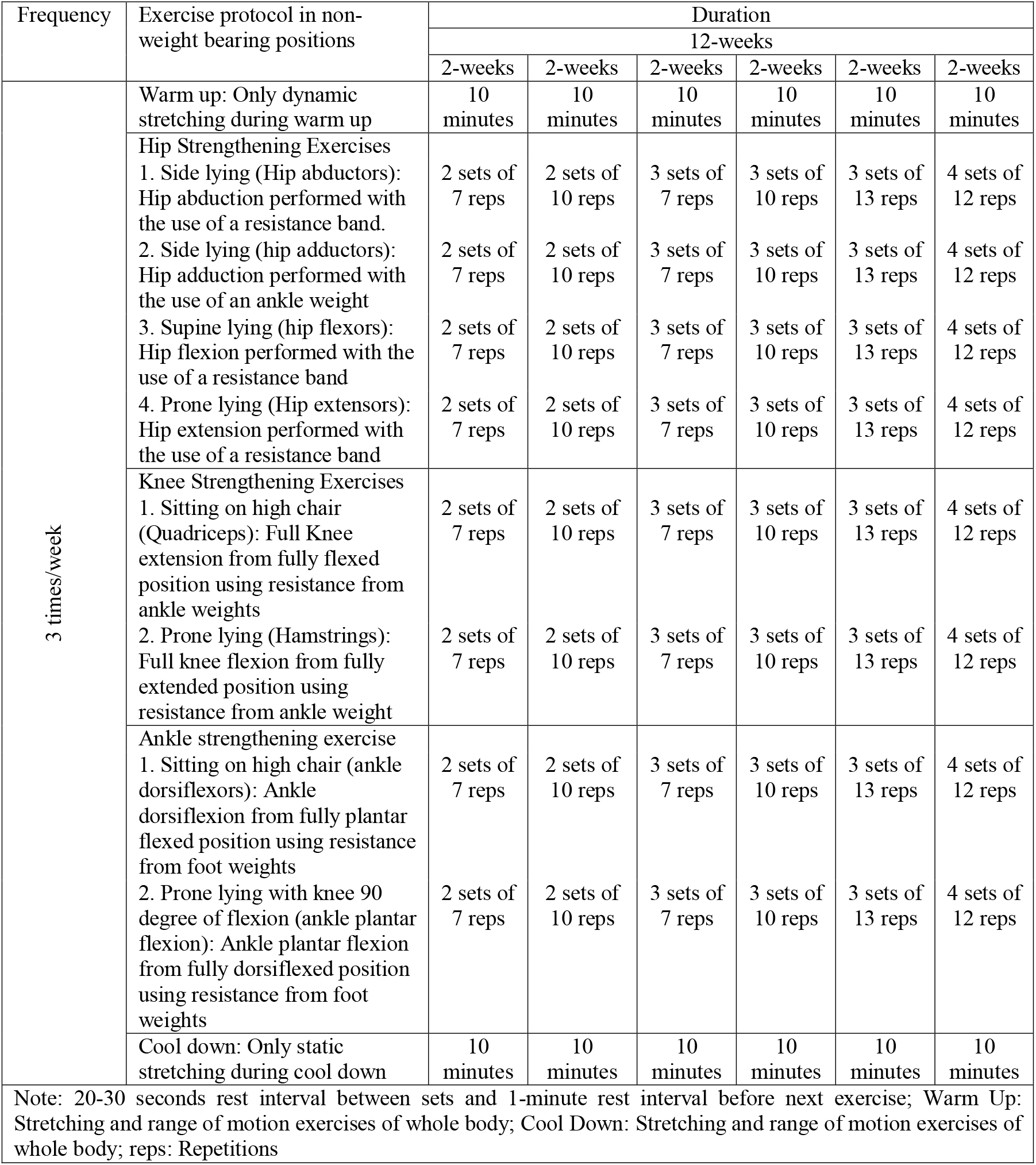
Lower Limb Rehabilitation Protocol (LLRP)

Each training session started with ten minutes of warm-up, forty-five to sixty minutes of lower limb strengthening exercises, and ten minutes of cool down at the end of the training protocol **(Table 1)**. The study demonstrated that dynamic stretching is recommended for warm-up to avoid a decrease in strength and performance (20). When the static stretching was used as part of a warm-up immediately prior to exercise, then it causes harm to muscle strength (21). The study explained that after two to four repetitions of static stretching, there is no increase in muscle elongation (22). A cool-down period is essential after a training session and should last approximately 5 to ten minutes (23, 24). Additionally, regular messages were also sent to the RGwo-mHealth for intention control.

#### Rehabilitation group with mobile health (RGw-mHealth)

Patients in the RGw-mHealth were prescribed with the LLRP as described in the RGwo-mHealth. Additionally, they receive regular reminders to carry out of LLRP using mHealth in the form of periodic manual WhatsApp messages. Two text messages per day for three days a week for a period of 12-weeks were sent to patients in the RGw-mHealth. Patients in this group received a total 72 text messages. The researcher sent text messages between 7:00 to 9:00 a.m. and 5:00 to 7:00 p.m. during the days of Wednesday, Friday, and Sunday. A study reported that sending text messages in the morning was to make sure that the patients have enough time to plan and do exercise during the day (25). The screen view for sending the text messages is shown in additional file 1.

#### Control group (CG)

Patients in the CG only followed the IDC for the duration of 12-weeks. The IDC contents include advice on general guidelines of mobility and healthy eating **(Table 2)**. The contents of IDC were translated into Urdu language by two language experts to ensure better patients’ understanding based on a recent pilot study (17). Regular messages were also sent to the CG for intention control.

**Table 2.** Instructions of daily care (IDC)

| Activity Name | IDC |
| --- | --- |
| Sitting | When there is option of sitting than standing, then prefer to sitting. Prefer your sitting on high stool or chair rather than low level. |
| Standing from Sitting | When you are standing from sitting position, and then initially sit at the edge of bed, chair or stool with the feet on the ground at the level of hips. Use the hands to push up from the bed, chair or stool. |
| Walking | Do not walk, jog or run as an exercise plan. Walking stick can be used on the opposite hand of the affected knee OA. If both knees are affected, then walker can be used. Use of knee brace and jogging shoes with well cushioned soles during walking is highly recommended. |
| Stair Climbing | Avoid stair climbing. But if there is need of stair climbing then support the side rails with your hands by placing the affected foot first on a stair step then the unaffected foot on the same step. |
| Working | Prefer working on a high stool or chair. |
| Body Weight | Try to reduce your weight by avoiding taking of sugary foods, drinks and high fat foods. Eat mostly plant-based foods. Add omega-3 fatty acids in your daily diet. |

IDC: Instructions of daily care

**Table 2.** Summary of the overall and baseline demographic characteristics and assessment scores of the QoL, functional strength and functional capacity of patients.

| **Demographic characteristics** | **Overall**<br>**(N = 114)** | **RGwo-**<br>**mHealth**<br>**(n = 38)** | **RGw-**<br>**mHealth**<br>**(n = 38)** | **CG**<br>**(n = 38)** |
| --- | --- | --- | --- | --- |
| **Age**, mean (SD), y | 53.00 (4.60) | 52.63 (4.60) | 53.97 (4.36) | 52.94 (4.58) |
| **Gender** (M/F) | 50/64 | 17/21 | 17/21 | 16/22 |
| **Socioeconomic status (according to Pakistan rupees), No. (%)** |  |  |  |  |
| Lower | 79 (69.3) | 28 (73.7) | 23 (60.5) | 28 (73.7) |
| Middle | 18 (15.8) | 7 (18.4) | 9 (23.7) | 2 (5.3) |
| Upper | 17 (14.9) | 3 (7.9) | 6 (15.8) | 8 (21.1) |
| **Employment, No. (%)** |  |  |  |  |
| Yes | 69 (60.5) | 24 (63.2) | 24 (63.2) | 21 (55.3) |
| No | 45 (39.5) | 14 (36.8) | 14 (36.8) | 17 (44.7) |
| **Weight (kg)**, mean (SD) | 87.05 (9.61) | 87.84 (9.42) | 89.47 (10.06) | 85.42 (9.29) |
| **BMI (Kg.m-2)**, mean (SD) | 32.17 (4.21) | 31.98 (3.61) | 33.06 (4.36) | 32.02 (4.65) |
| **Quality of life by WOMAC summary, mean (SD)** | 41.59 (12.71) | 40.65 (13.49) | 45.44 (12.13) | 38.05 (12.08) |
| **Functional strength by FRSST (seconds)** | 19.40 (4.21) | 19.50 (3.41) | 21.72 (4.38) | 16.98 (3.45) |
| **Functional capacity by GST (cm/s), mean (SD)** | 63.25 (7.28) | 64.32 (7.31) | 62.13 (8.34) | 63.29 (6.04) |

Abbreviations: BMI = Body mass index; M = Male; F = Female; WOMAC = Western Ontario and McMaster Universities Osteoarthritis Index; FRSST = Five repetitions sit-to-stand test; GST = Gait speed test; SD = Standard Deviation; N = Number

#### Measurements

Patient’s demographics, QoL, functional strength, and functional capacity were assessed at baseline before randomization. Similarly, the assessments of demographics, QoL, functional strength, and functional capacity were repeated at posttest after 12 weeks of intervention. The demographic questionnaire covered age, gender, socioeconomic status, and employment. Outcome measures were categorized into primary and secondary outcome measures.

#### Primary Outcome Measures

Primary outcome measures were QoL and functional strength. To evaluate QoL, the Western Ontario and McMaster Universities Osteoarthritis Index (WOMAC) summary score that was already adapted and validated was used (26). Three dimensional (pain, stiffness, and physical function) QoL questionnaire was designed for the evaluation of the hip and knee OA. The WOMAC score ranges from 0 to 4 on a Likert-type scale, with higher scores indicating an increase in pain, joint stiffness, and reduced functionality (27).

The FRSST was used for the assessment of lower body functional strength. The FRSST has a high test-retest reliability for adults and subjects with knee OA (28, 29). A straight armless chair with 43 cm of back support height was stabilized on the wall. Patients were asked to come forward on the chair seat in a position of standing, until the feet are flat on the floor. Patients were instructed to stand up and sit down once without using the upper limbs as a test purpose. For those who needed assistance, assistance was provided to complete the test. Patients were then asked to stand up and sit down as quickly as possible five times. Timing with a stopwatch was started on the command go and ceased when the patient completed the 5th set of sit to stand. The time taken was recorded as the patients’ score.

#### Secondary outcome measures

Secondary outcome measure was the functional capacity. The Gait Speed Test (GST) was used for the assessment of the functional capacity. In the GST, the time was recorded when the patient completed a distance of 20 feet and then divided the distance to time for the calculation of gait speed. Gait speed measures obtained during a single test session are reliable. The coefficients (0.90) of comfortable gait speed were highly reliable (30). The GST is used as an outcome measure in rehabilitation (31) and in trials of interventions to delay the onset of disability or frailty (32).

#### Statistical procedures

The Statistical Package for Social Sciences, version 22, Chicago, IL, was used to analyze the data. Continuous variables were presented as mean (standard deviation [SD]) based on data distribution. The Shapiro-Wilk test was used to assess the normality of all variables. Categorical variables were presented as frequencies (n) and percentages (%). For categorical demographic variables, the One-Way Analysis of Variance (ANOVA) was used to compare for differences between variables. Since all data was normally distributed; the Paired Samples t-test was used to analyze differences between the baseline and post-test assessments within the groups.

The overall treatment effects on change in clinical outcome measures were estimated using the One Way ANOVA (unadjusted results) and Analyses of Covariance (ANCOVA, adjusted results) for mean changes (95% confidence interval [CI]) from baseline to posttest assessments in the continuous outcome data. The ANCOVA model included the changes as the dependent variable, with group as a main effect and the baseline scores as an additional covariate. The purpose of using the pretest (baseline) scores as a covariate in ANCOVA with a pretest-posttest design is to reduce the error variance and eliminate systematic bias [33]. The pairwise comparisons between groups were estimated using Bonferroni post hoc test. The value of p ≤ 0.05 was considered statistically significant.

## RESULTS

Figure 1 shows a patient flow chart in the current study. A total of 156 overweight and obese knee OA patients were initially enrolled and assessed for eligibility. Among them, 42 patients were excluded for reasons as shown in Figure 1. The remaining 114 were randomized equally into three groups. The post-intervention outcomes were not obtained for the 18 withdrawn patients. Figure 1 demonstrates the study flow chart including reasons given by patients who did not complete the study. A final total of 96 patients (32 in the RGw-mHealth, 32 in the RGwo-mHealth, and 32 in the CG) were included in the analysis of QoL, functional strength and functional capacity.

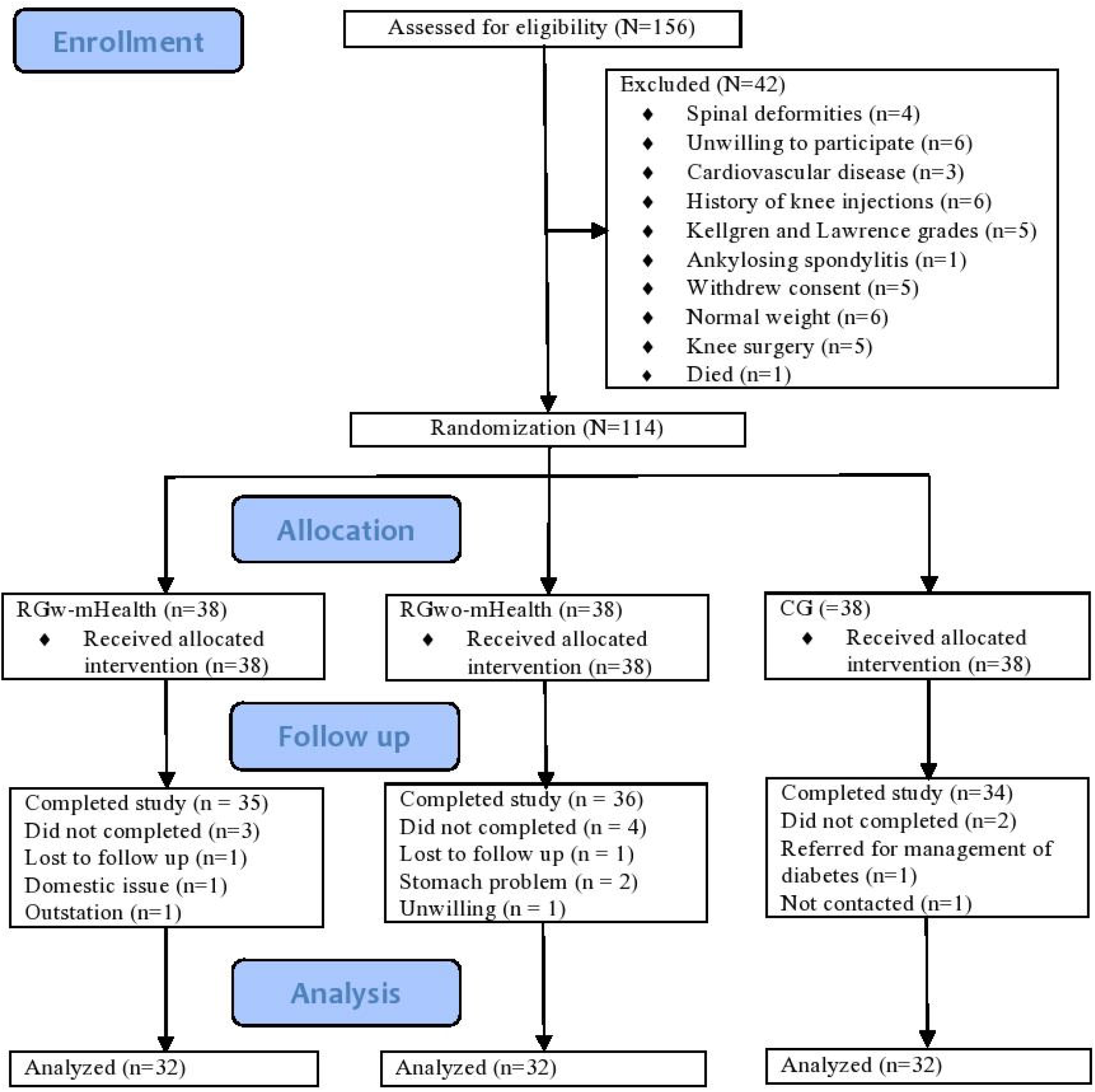

Non-completers did not differ significantly from completers in terms of age, gender, socioeconomic status, employment, QoL, functional strength, and functional capacity. Similarly, retention did not differ among the groups (RGw-mHealth, 84%; RGwo-mHealth, 84%; CG, 84%). Two serious adverse events (one appendix surgery in the RGw-mHealth and one gallbladder surgery in the RGwo-mHealth) were unrelated to the study. One nonserious adverse event of muscle spasm in the CG was noted during the study and it was related to the study.

Summary of the overall and baseline demographic characteristics and assessment scores of the QoL, functional strength, and functional capacity are described in **Table 3**. After participation in the 12-weeks of intervention, a statistically significant improvement compared to baseline was observed for QoL, functional strength, and functional capacity scores in the RGw-mHealth and RGwo-mHealth. In the CG, improvement in QoL score was also statistically significant **(Table 4)**.

**Table 3.** Change in clinical outcome measures from baseline to post-test after 12-weeks of interventions within groups.

| **Outcome measures** | **Group** | **Baseline, Mean (SD)** | **Post-test, Mean (SD)** | **Change, Mean (95% CI)** | **p-value** |
| --- | --- | --- | --- | --- | --- |
| **QoL by WOMAC Summary (0-96)** | RGwo-mHealth | 40.53 (13.71) | 36.17 (13.02) | 4.35 (3.09, 5.62) | <0.001 |
|  | RGw-mHealth | 45.81 (11.65) | 36.53 (11.84) | 9.28 (8.77,9.78) | <0.001 |
|  | CG | 37.15 (12.28) | 36.40 (12.08) | 0.75 (0.25, 1.24) | 0.004 |
| **Functional strength by FRSST (seconds)** | RGwo-mHealth | 19.90 (3.48) | 16.87 (2.62) | 3.03 (1.99, 4.07) | <0.001 |
|  | RGw-mHealth | 21.43 (4.11) | 16.00 (3.27) | 5.43 (4.37, 6.49) | <0.001 |
|  | CG | 17.20 (3.29) | 16.67 (3.08) | 0.52 (-0.01, 1.05) | 0.056 |
| **Functional capacity by GST (cm/s)** | RGwo-mHealth | 63.36 (7.32) | 72.42 (11.12) | -9.06 (-11.60, -6.51) | <0.001 |
|  | RGw-mHealth | 60.81 (8.22) | 84.46 (13.88) | -23.65 (-27.56, -19.74) | <0.001 |
|  | CG | 62.70 (6.24) | 63.88 (6.81) | -1.18 (-2.46, 0.09) | 0.067 |

Abbreviations: WOMAC = Western Ontario and McMaster Universities Osteoarthritis Index; CI = Confidence interval; FRSST = Five repetitions sit-to-stand test; GST = Gait speed test

**Table 3.** Summary of the overall and baseline demographic characteristics and assessment scores of the QoL, functional strength and functional capacity of patients

| Demographic characteristics | Overall<br>(N = 114) | RGwo-mHealth<br>(n = 38) | RGw-mHealth<br>(n = 38) | CG<br>(n = 38) |
| --- | --- | --- | --- | --- |
| Age, mean (SD), y | 53.00 (4.60) | 52.63 (4.60) | 53.97 (4.36) | 52.94 (4.58) |
| Gender |  |  |  |  |
| • Male | 50 | 17 | 17 | 16 |
| • Female | 64 | 21 | 21 | 22 |
| Socioeconomic status (according to<br>Pakistan rupees), No. (%) |  |  |  |  |
| • Lower | 79 (69.3) | 28 (73.7) | 23 (60.5) | 28 (73.7) |
| • Middle | 18 (15.8) | 7 (18.4) | 9 (23.7) | 2 (5.3) |
| • Upper | 17 (14.9) | 3 (7.9) | 6 (15.8) | 8 (21.1) |
| Employment, No. (%) |  |  |  |  |
| • Yes | 69 (60.5) | 24 (63.2) | 24 (63.2) | 21 (55.3) |
| • No | 45 (39.5) | 14 (36.8) | 14 (36.8) | 17 (44.7) |
| Weight (kg), mean (SD) | 87.05 (9.61) | 87.84 (9.42) | 89.47 (10.06) | 85.42 (9.29) |
| BMI (Kg.m-2), mean (SD) | 32.17 (4.21) | 31.98 (3.61) | 33.06 (4.36) | 32.02 (4.65) |
| Quality of life by WOMAC<br>summary, mean (SD) | 41.59 (12.71) | 40.65 (13.49) | 45.44 (12.13) | 38.05 (12.08) |
| Functional strength by FRSST<br>(seconds) | 19.40 (4.21) | 19.50 (3.41) | 21.72 (4.38) | 16.98 (3.45) |
| Functional capacity by GST (cm/s),<br>mean (SD) | 63.25 (7.28) | 64.32 (7.31) | 62.13 (8.34) | 63.29 (6.04) |

BMI: Body mass index; M: Male; F: Female; WOMAC: Western Ontario and McMaster Universities Osteoarthritis Index; FRSST: Five repetitions sit-to-stand test; GST: Gait speed test; SD: Standard Deviation; N: Number

**Table 4.** Unadjusted and adjusted treatment effects on change in clinical outcome measures from baseline at post-test after 12 weeks of interventions and significance between groups.

| Change in variables | Unadjusted or adjusted | RGwo-mHealth (n = 32)<br>Mean (95% CI) | RGw-mHealth (n = 32)<br>Mean (95% CI) | CG (n = 32)<br>Mean (95% CI) | p-value |
| --- | --- | --- | --- | --- | --- |
| **Δ QoL by WOMAC Summary, (0-96)** | Unadjusted | 4.35 (3.09, 5.62) | 9.28 (8.77, 9.78) | 0.75 (0.25, 1.24 ) | <0.001 |
|  | Adjusted | 4.38 (3.57, 5.18) | 9.11 (8.28, 9.93) | 0.89 (0.08, 1.71) | <0.001 |
| **Δ Functional strength by FRSST, (seconds)** | Unadjusted | 3.03 (1.99, 4.07) | 5.43 (4.37, 6.49) | 0.52 (-0.01, 1.05) | <0.001 |
|  | Adjusted | 2.87 (2.14, 3.60) | 4.66 (3.90, 5.43) | 1.44 (0.67, 2.22) | <0.001 |
| **Δ Functional capacity by GST (cm/s)** | Unadjusted | -9.06 (-11.60, -6.51) | -23.65 (-27.56, -19.74) | -1.18 (-2.46, 0.09) | <0.001 |
|  | Adjusted | -8.98 (-11.72, -6.24) | -23.75 (-26.50, -21.01) | -1.15 (-3.88, 1.57) | <0.001 |

Abbreviations: WOMAC = Western Ontario and McMaster Universities Osteoarthritis Index; CI = Confidence interval; FRSST = Five repetitions sit-to-stand test; GST = Gait speed test

**Table 4.** Change in clinical outcome measures from baseline to post-test after 12-weeks of interventions within groups

| Outcome measures | Group | Baseline, Mean (SD) | Post-test, Mean (SD) | Change, Mean (95% CI) | *p* |
| --- | --- | --- | --- | --- | --- |
| QoL by WOMAC Summary (0-96) | RGwo-mHealth | 40.53 (13.71) | 36.17 (13.02) | 4.35 (3.09, 5.62) | <0.001 |
|  | RGw-mHealth | 45.81 (11.65) | 36.53 (11.84) | 9.28 (8.77, 9.78) | <0.001 |
|  | CG | 37.15 (12.28) | 36.40 (12.08) | 0.75 (0.25, 1.24) | 0.004 |
| Functional strength by FRSST (seconds) | RGwo-mHealth | 19.90 (3.48) | 16.87 (2.62) | 3.03 (1.99, 4.07) | <0.001 |
|  | RGw-mHealth | 21.43 (4.11) | 16.00 (3.27) | 5.43 (4.37, 6.49) | <0.001 |
|  | CG | 17.20 (3.29) | 16.67 (3.08) | 0.52 (-0.01, 1.05) | 0.056 |
| Functional capacity by GST (cm/s) | RGwo-mHealth | 63.36 (7.32) | 72.42 (11.12) | -9.06 (-11.60, -6.51) | <0.001 |
|  | RGw-mHealth | 60.81 (8.22) | 84.46 (13.88) | -23.65 (-27.56, -19.74) | <0.001 |
|  | CG | 62.70 (6.24) | 63.88 (6.81) | -1.18 (-2.46, 0.09) | 0.067 |

WOMAC: Western Ontario and McMaster Universities Osteoarthritis Index; CI: Confidence interval; FRSST: Five repetitions sit-to-stand test; GST: Gait speed test

The mean changes in QoL scores were 9.11 (95% CI 8.28, 9.93), 4.38 (95% CI 3.57, 5.18), and 0.89 (95% CI 0.08, 1.71) for patients in the RGw-mHealth, RGwo-mHealth and CG respectively (**Table 5)**. The pairwise between-group comparisons of the QoL score at post-test revealed that patients in the RGw-mHealth had significantly better QoL scores relative to both the RGwo-mHealth (difference, 4.72 [3.31, 6.14]; p < 0.001) and CG (difference, 8.21 [6.76, 9.66]; p < 0.001). There was also a statistically significant difference in the mean QoL scores between the RGwo-mHealth and CG (difference, 3.48 [2.08, 4.88]; p < 0.001) **(Table 6)**.

**Table 5.**
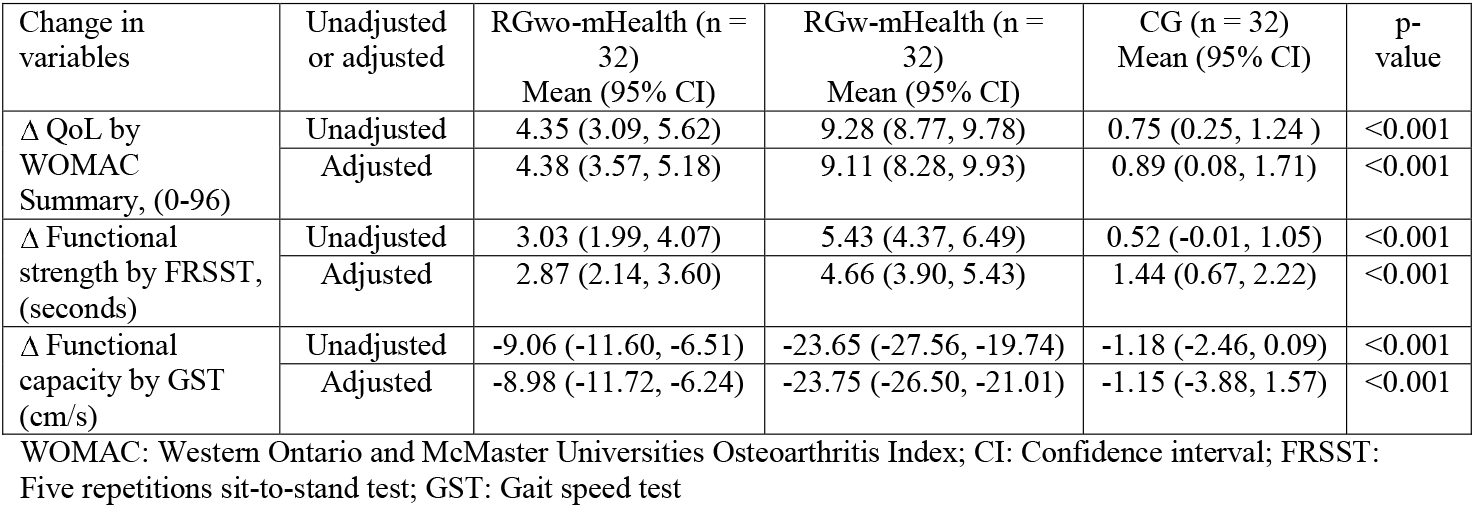
Unadjusted and adjusted treatment effects on change in clinical outcome measures from baseline at post-test after 12 weeks of interventions and significance between groups

**Table 6.** The pairwise comparisons between groups by Bonferroni test (post hoc test)

| Variables | Comparison between Groups | Mean difference (95 % CI for difference) | *p* |
| --- | --- | --- | --- |
| Quality of life by WOMAC Summary, (0-96) | RGw-mHealth and RGwo-mHealth | 4.72 (3.31, 6.14) | *<0.001* |
|  | RGwo-mHealth and CG | 3.48 (2.08, 4.88) | *<0.001* |
|  | RGw-mHealth and CG | 8.21 (6.76, 9.66) | *<0.001* |
| Functional strength by FRSST (seconds) | RGw-mHealth, RGwo-mHealth | -1.79 (-3.07, -0.51) | *0.003* |
|  | RGwo-mHealth and CG | 1.42 (0.10, 2.74) | *0.030* |
|  | RGw-mHealth and CG | 3.22 (1.81, 4.62) | *<0.001* |
| Functional capacity by GST, (cm/s) | RGw-mHealth, RGwo-mHealth | 14.77 (9.98, 19.56) | *<0.001* |
|  | RGwo-mHealth and CG | -7.82 (-12.56, -3.08) | *<0.001* |
|  | RGw-mHealth and CG | -22.60 (-27.36, -17.835) | *<0.001* |

RGw-mHealth: Rehabilitation group with mobile health; RGwo-mHealth: Rehabilitation group without mobile health; CG: Control group; WOMAC: Western Ontario and McMaster Universities Osteoarthritis Index; CI: Confidence interval; FRSST: Five repetitions sit-to-stand test; GST = Gait speed test

**Table 5** presents the mean change in the FRSST (seconds), as an indication of the participant’s functional strength change after 12-weeks. The mean change in FRSST scores in the RGw-mHealth, RGwo-mHealth, and CG were 4.66 seconds [95% CI 3.90, 5.43 seconds], 2.87 seconds [95% CI 2.14, 3.60 seconds] and 1.44 seconds [95% CI 0.67, 2.22 seconds] respectively. As indicated by the overall ANCOVA, there was a statistically significant change in functional strength between groups (p < 0.001) after 12 weeks of intervention. The pairwise between-group comparisons of the mean change in functional strength at post-test revealed that patients in the RGw-mHealth had statistically significant greater mean changes in functional strength relative to both the RGwo-mHealth (difference, -1.79 [-3.07, -0.51]; p = 0.003) and CG (difference, 3.22 [1.81, 4.62]; p < 0.001). There was also a statistically significant difference in functional strength between the RGwo-mHealth and CG (difference 1.42 [0.10, 2.74]; p = 0.030) as shown in **Table 6**.

Patients in the RGw-mHealth had a mean change in gait speed of -23.75 (95% CI - 26.50, -21.01) at post-test, whereas patients in the RGwo-mHealth and CG demonstrated a gait speed mean change of -8.98 (95% CI -11.72, -6.24) and -1.15 (95% CI -3.88, 1.57) respectively as shown in **Table 5**. The pairwise between-group comparisons of functional capacity at posttest revealed that patients in the RGw-mHealth had a significantly larger mean changes in functional capacity score compared to both the RGwo-mHealth (difference, 14.77 [9.98, 19.56]; p < 0.001) and CG (difference, -22.60 [-27.36, -17.835]; p < 0.001). There was also a statistically significant difference in the mean change in functional capacity between RGwo-mHealth and CG (difference, -7.82 [-12.56, -3.08]; p < 0.001) as shown in **Table 6**.

## DISCUSSION

The core recommended treatments for OA in OA clinical guidelines are lower limb muscle strengthening (34). Exercise is often indicated as one of the main components in the rehabilitation process (35). Effectiveness of rehabilitation combined with mHealth may provide more objective data than the standard rehabilitation approaches we are using today to treat overweight and obese knee OA patients. Therefore, the current study investigated the effectiveness of the LLRP using mHealth on QoL, functional strength, and functional capacity among knee OA patients who were overweight and obese. We reached the conclusion that in RGw-mHealth patients, the scores of QoL, functional strength, and functional capacity were significantly higher than RGwo-mHealth or CG. The results indicated that patients in the RGw-mHealth who received additional reminders of the intervention combined with WhatsApp messages showed greater improvements in QoL, functional strength, and functional capacity than did the patients in the RGwo-mHealth or CG. In the current study, knee OA was more common in females, while there was no difference in terms of age. The mean age of the overall sample was 53.00 years (SD = 4.60 years). The majority of the study patients were employed (60.5%), and had lower socioeconomic status (69.3%).

In a randomized clinical trial of 316 overweight or obese elderly men and women with knee OA, it was noted that the combination of diet and exercise was more successful in improving health-related QoL, if compared with exercise or diet as single interventions (36). Because there is no cure for this condition, current medical practice focuses on such interventions that reduce the progression of the disease and the negative impact on health-related QoL (37). A recent study concluded that the progressive resistance strength training of LLRP is effective in improving QoL among overweight and obese knee OA patients (38). In the current study, the patients in the RGw-mHealth resulted significantly better QoL compared to both the RGwo-mHealth and CG.

Recently, a study demonstrated that a combination of dietary weight loss and exercise intervention was consistently better in improving a combination of performance and functional outcomes among participants with knee OA compared with exercise alone, diet alone, or a control group (39). In a recent study, a combination of IDC with the strengthening exercises of the major muscle groups of the lower limbs in non-weight-bearing positions resulted in improving functional capacity in overweight and obese knee OA patients (38). Similarly, in the current study the intervention in the patients of RGw-mHealth and RGwo-mHealth was also a combination of IDC and progressive LLRP that reported significant results in improving functional capacity. According to the Osteoarthritis Research Society International Committee for Clinical Trials Response Criteria Initiative and the Outcome Measures in Rheumatology Committee, response to treatment in clinical trials should be based on symptomatic response to therapy in the domain of function, and the patient’s global assessment (40). Data from a study involving knee OA patients who were obese highlight an opportunity to improve the QoL scores by following nutritional education and dietary guidelines (41). In the current study, the IDC also focused on the caloric restriction diet.

Many trials of different physical activity and exercise-based interventions reported the improvement of function among knee OA patients (42). Patients in the RGw-mHealth had statistically significant greater functional strength compared to both the RGwo-mHealth and CG. This may be due to the strengthening exercises of LLRP that were performed in non-weight-bearing positions by patients in the RGw-mHealth in the current study. These strengthening exercises resulted in increased strength of the lower limb muscles. Therefore, the patients in the RGw-mHealth had better improvement in the score of functional strength than the patients in the RGwo-mHealth and CG.

In a meta-analysis, it was indicated that the number of directly supervised exercise sessions can influence the extent of the treatment effect (43). In the current study, the strengthening exercise sessions in the RGw-mHealth were performed 3 times a week for 12 weeks (36 sessions) by sending periodic manual WhatsApp messages as a reminder to perform their intervention. Therefore, the treatment effects of patients in the RGw-mHealth were better than the patients in the RGwo-mHealth and CG.

Based on these findings, the strengthening exercises of LLRP using mHealth are expected to be more effective in terms of improving functional strength, functional capacity, and QoL than any other rehabilitation intervention among overweight and obese knee OA patients. The strengthening exercises of LLRP by using mHealth may have the potential to improve QoL, functional strength, and functional capacity among overweight and obese knee OA patients. These exercises may have the great contribution to the body of knowledge internationally because these are performed in non-weight bearing sitting or lying positions with minimal load at the knee joint. In addition, this rehabilitation protocol is easy to use in the home care setting and may be helpful in treating many bedridden conditions such as multiple sclerosis, hemiplegia, paraplegia and neurological diseases with lower limb weakness.

The present study had some limitations. It was conducted in a single centre to recruit patients. No long-term follow-up records were taken. Thus, further blinded studies across multiple centres and long-term follow-up are required to confirm the results of the strengthening exercises of the LLRP combined with mHealth in overweight and obese knee OA patients. Additionally, physical activity, psychosocial, and comorbidity factors may influence the outcomes. Therefore, further research considering these additional factors is required to confirm the findings of the study.

In conclusion, patients in the RGw-mHealth had better improvement in functional strength, functional capacity, and QoL than those in the RGwo-mHealth and CG. The results of the current study suggest that improvements in functional strength, functional capacity, and QoL among knee OA patients who are overweight or obese, are augmented better by the implementation of the LLRP using mHealth to rehabilitation or general treatment without mHealth.

## Data Availability

All data produced in the present study are available upon reasonable request to the authors

## Data Availability

The data used to support the findings of the study are available from the corresponding author upon request.

## REFERENCES

1. Barbour KE, Helmick CG, Boring M, Zhang X, Lu H, Holt JB. Prevalence of doctor-diagnosed arthritis at state and county levels—United States, 2014. Morbidity and Mortality Weekly Report 2016;65(19):489–94.

2. Taruc-Uy RL, Lynch SA. Diagnosis and treatment of osteoarthritis. Primary Care: Clin Office Pract 2013;40(4):821–36.

3. Helmark IC, Mikkelsen UR, Børglum J, Rothe A, Petersen MC, Andersen O, Langberg H, Kjaer M. Exercise increases interleukin-10 levels both intraarticularly and peri-synovially in patients with knee osteoarthritis: a randomized controlled trial. Arthritis Res Ther 2010;12(4):R126.

4. Bridges SL. National institute of arthritis and musculoskeletal and skin diseases. Arthritis Res Ther 2000;2(1):0003.

5. Godin G, Desharnais R, Valois P, Lepage L, Jobin J, Bradet R. Differences in perceived barriers to exercise between high and low intenders: observations among different populations. Am J Health Promot 1994;8(4):279–85.

6. Reichert FF, Barros AJ, Domingues MR, Hallal PC. The role of perceived personal barriers to engagement in leisure-time physical activity. Am J Public Health 2007;97(3):515–9.

7. Maetzel A, Li LC, Pencharz J, Tomlinson G, Bombardier C; Community Hypertension and Arthritis Project Study Team. The economic burden associated with osteoarthritis, rheumatoid arthritis, and hypertension: a comparative study. Ann Rheum Dis 2004;63(4):395–401.

8. Vos T, Allen C, Arora M, Barber RM, Bhutta ZA, Brown A, et al. Global, regional, and national incidence, prevalence, and years lived with disability for 310 diseases and injuries, 1990–2015: a systematic analysis for the Global Burden of Disease Study 2015. The Lancet 2016;388(10053):1545–602.

9. Lawrence RC, Felson DT, Helmick CG, Arnold LM, Choi H, Deyo RA, et al. Estimates of the prevalence of arthritis and other rheumatic conditions in the United States: Part II. Arthritis Rheum 2008;58(1):26–35.

10. Bohannon RW. Sit-to-stand test for measuring performance of lower extremity muscles. Percept Mot Skills 1995 Feb;80(1):163–6.

11. Wang TJ. Concept analysis of functional status. Int J Nurs Stud 2004 May;41(4):457–62.

12. Wantland DJ, Portillo CJ, Holzemer WL, Slaughter R, McGhee EM. The effectiveness of Web-based vs. non-Web-based interventions: a meta-analysis of behavioral change outcomes. J Med Internet Res 2004;6(4):e40.

13. Han M, Lee E. Effectiveness of mobile health application use to improve health behavior changes: a systematic review of randomized controlled trials. Healthcare informatics research 2018;24(3):207–26.

14. Fransen M, McConnell S, Harmer AR, Van der Esch M, Simic M, Bennell KL. Exercise for osteoarthritis of the knee: a Cochrane systematic review. Br J Sports Med 2015; 49(24):1554–7.

15. Ottawa Panel Members, Ottawa Methods Group, Brosseau L, Wells GA, Tugwell P, Egan M, Dubouloz CJ, Casimiro L, et al. Ottawa panel evidence-based clinical practice guidelines for therapeutic exercises and manual therapy in the management of osteoarthritis. Phys Ther 2005 Sep 1;85(9):907–71.

16. Rafiq MT A, Hamid MS, Hafiz E. Non-pharmacological interventions for treating symptoms of knee osteoarthritis in overweight or obese patients; A review. J Postgrad Med Inst 2020; 34(3): 142–8.

17. Rafiq, M. T. A, Hamid, M. S., Eliza, H. A., & Sakib A. Rehabilitation protocol with or without mobile health in overweight and obese knee osteoarthritis patients-a pilot study. Balneo Research Journal 2019;10(4):580–584. doi:10.12680/balneo.2019.306

18. WHO Expert Consultation. Appropriate body-mass index for Asian populations and its implications for policy and intervention strategies. Lancet 2004 Jan 10;363(9403):157–63. doi: 10.1016/S0140-6736(03)15268-3.

19. Kellgren JH, Lawrence JS. Radiological assessment of osteo-arthrosis. Ann Rheum Dis 1957;16(4):494.

20. Page P. Current concepts in muscle stretching for exercise and rehabilitation. Int J Sports Phys Ther 2012 Feb;7(1):109–19.

21. McHugh MP, Nesse M. Effect of stretching on strength loss and pain after eccentric exercise. Medicine and science in sports and exercise 2008 Mar 1;40(3):566.

22. Taylor DC, Dalton JR JD, Seaber AV, Garrett JR WE. Viscoelastic properties of muscle-tendon units: the biomechanical effects of stretching. Am J Sports Med1990 May;18(3):300–9.

23. Powers SK, Howley ET, Quindry J. Exercise physiology: Theory and application to fitness and performance. New York, NY: McGraw-Hill; 2007.

24. Prentice WE. The thigh, hip, groin, and pelvis. Arnheim’s principles of athletic training: A competency-based approach 2003:656–98.

25. Gell NM, Wadsworth DD. The use of text messaging to promote physical activity in working women: A randomized controlled trial. J Phys Act Health 2015 Jun 1;12(6):756–63.

26. Alexandre TD, Cordeiro RC, Ramos LR. Factors associated to quality of life in active elderly. Revista de saude publica 2009;43:613–21.

27. Grotle M, Hagen KB, Natvig B, Dahl FA, Kvien TK. Obesity and osteoarthritis in knee, hip and/or hand: an epidemiological study in the general population with 10 years follow-up. BMC Musculoskelet Disord 2008;9(1):132.

28. Bohannon RW. Test-retest reliability of the five-repetition sit-to-stand test: a systematic review of the literature involving adults. J Strength Cond Res 2011;25(11):3205–7.

29. Schlenk EA, Lias JL, Sereika SM, Dunbar□acob J, Kwoh CK. Improving physical activity and function in overweight and obese older adults with osteoarthritis of the knee: a feasibility study. Rehabil Nurs 2011;36(1):32–42.

30. Bohannon RW. Comfortable and maximum walking speed of adults aged 20—79 years: reference values and determinants. Age and ageing 1997;26(1):15–9.

31. Skinner A, Turner-Stokes L. The use of standardized outcome measures in rehabilitation centres in the UK. Clin Rehabil 2006 Jul;20(7):609–15.

32. Fairhall N, Aggar C, Kurrle SE, Sherrington C, Lord S, Lockwood K, Monaghan N, Cameron ID. Frailty intervention trial (FIT). BMC geriatrics 2008 Dec;8(1):1–0.

33. Dimitrov DM, Rumrill Jr PD. Pretest-posttest designs and measurement of change. Work 2003 Jan 1;20(2):159–65.

34. Heidari B. Knee osteoarthritis prevalence, risk factors, pathogenesis and features: Part I. Caspian J Intern Med 2011;2(2):205.

35. Bartholdy C, Juhl C, Christensen R, Lund H, Zhang W, Henriksen M. The role of muscle strengthening in exercise therapy for knee osteoarthritis: A systematic review and meta-regression analysis of randomized trials. Semin Arthritis Rheum 2017 Aug;47(1):9–21.

36. Rejeski WJ, Focht BC, Messier SP, Morgan T, Pahor M, Penninx B. Obese, older adults with knee osteoarthritis: weight loss, exercise, and quality of life. Health Psychol 2002 Sep;21(5):419.

37. Rejeski WJ, Brawley LR, Shumaker SA. Physical activity and health-related quality of life. Exerc Sport Sci Rev 1996 Jan 1;24(1):71–108.

38. Rafiq MT A, Hamid MS, Hafiz E. Effect of Progressive Resistance Strength Training on Body Mass Index, Quality of Life and Functional Capacity in Knee Osteoarthritis: A Randomized Controlled Trial. J Multidiscip Healthc 2021;14:2161–2168. Published 2021 Aug 11. doi:10.2147/JMDH.S317896

39. Messier SP, Loeser RF, Miller GD, Morgan TM, Rejeski WJ, Sevick MA, Ettinger Jr WH, Pahor M, Williamson JD. Exercise and dietary weight loss in overweight and obese older adults with knee osteoarthritis: the Arthritis, Diet, and Activity Promotion Trial. Arthritis Rheum 2004 May;50(5):1501–10.

40. Pham T, van der Heijde DM, Altman RD, Anderson JJ, Bellamy N, Hochberg M, Simon L, Strand V, Woodworth T, Dougados M. OMERACT-OARSI initiative: Osteoarthritis Research Society International set of responder criteria for osteoarthritis clinical trials revisited. Osteoarthritis Cartilage 2004 May 1;12(5):389–99.

41. Gomes-Neto M, Araujo AD, Junqueira ID, Oliveira D, Brasileiro A, Arcanjo FL. Comparative study of functional capacity and quality of life among obese and nonobese elderly people with knee osteoarthritis. Revista Brasileira de Reumatologia (English Edition) 2016 Mar 1;56(2):126–30.

42. Verhagen AP, Ferreira M, Reijneveld-van de Vendel EA, Teirlinck CH, Runhaar J, van Middelkoop M, Hermsen L, de Groot IB, Bierma-Zeinstra SM. Do we need another trial on exercise in patients with knee osteoarthritis? no new trials on exercise in knee oa. Osteoarthritis Cartilage 2019 Sep 1;27(9):1266–9.

43. Fransen M, McConnell S. Exercise for osteoarthritis of the knee. Cochrane Database Syst Rev 2008 Oct 8;(4):CD004376.

