## Supplementary Files 1 for "Effectiveness of lower limb rehabilitation protocol using mobile health on quality of life, functional strength and functional capacity among knee osteoarthritis patients who were overweight and obese: A randomized controlled trial"

**Additional file 1:** Screen View for Sending the Text Messages

Assalamualaikum,

This is reminder to carry out the LLRP and IDC.

Your week/weeks………of training is going on.

Please follow it for a better life.

Your Caring Doctor

Muhammad Tariq Rafiq
